# Intracranial Stimulation in Epilepsy as a Novel Approach for Mapping Psychiatric Circuits

**DOI:** 10.1101/2025.06.28.25330302

**Authors:** Haun Sun, Ningfei Li, Alejandro Granados Martinez, Fahmida A. Chowdhury, Beate Diehl, Harith Akram, Veerle Visser-Vandewalle, Bryan Strange, Juan A. Barcia, Mircea Polosan, Stéphan Chabardes, Davide Giampiccolo, Vladimir Litvak

## Abstract

**Objective:** To assess the feasibility of using stereo-electroencephalography (SEEG) depth electrodes, implanted for invasive recording of the epileptogenic zone in drug-resistant epilepsy, to identify and validate novel Deep Brain Stimulation (DBS) targets for psychiatric disorders.

**Methods:** SEEG data from 86 patients with drug-resistant epilepsy implanted at the UK National Hospital for Neurology and Neurosurgery were analysed (6,372 electrode contacts). Normative structural and functional connectomes were combined with meta-analytic functional maps to estimate the probability of each contact engaging networks associated with psychiatric symptoms. A linear model, developed using published data from 80 patients with obsessive-compulsive disorder (OCD) treated with DBS, was used to validate the approach.

**Results:** Networks involved in reward processing emerged as the most promising DBS targets, with a predicted required sample size of 7 patients (range 4–33). Bipolar disorder was the next most feasible (22 patients; range 10–44), followed by depression (32; range 16–49) and anxiety (38; range 17–59). For other disorders, estimated sample sizes exceeded 40, indicating limited feasibility for single-centre studies.

**Conclusions:** This study provides preliminary support for using SEEG to map brain circuits underlying psychiatric symptoms. The method leverages routinely implanted electrodes in epilepsy to explore candidate DBS targets, supporting a network-guided strategy for personalised neuromodulation in neuropsychiatry.

## Introduction

Brain stimulation refers to the direct application of electromagnetic energy to the brain to modulate pathological neural circuits. Among available techniques, Deep Brain Stimulation (DBS)—a chronic, invasive method targeting deep brain regions—has proven effective for various neurological and psychiatric disorders. The modern version of DBS was initially developed in the late 1980s for Parkinson’s disease, and its applications have steadily expanded since [1]. Recent research focuses on treating severe, medication-resistant psychiatric conditions, such as major depressive disorder and obsessive-compulsive disorder (OCD). Another significant advancement in the field is the integration of normative connectomes with analyses of DBS effects [2]. This approach aims to identify functional and structural brain networks that should be targeted to achieve therapeutic benefits (‘sweet spots’) or avoided to prevent side effects (’sour spots’) [3–5]. Similar techniques are also employed for network mapping based on brain lesion data and non-invasive stimulation effects [6]. Despite these advances, evaluating DBS effects on novel targets in humans remains challenging.

Potential insights could come from data acquired in patients with drug-resistant focal epilepsy who undergo stereotactic EEG (SEEG) recordings as part of their standard preoperative evaluation for potential resection surgery [7–9] These patients are implanted according to their individual seizure semiology and suspected epileptogenic network arising from dysfunction in a region in the cortex or medial temporal lobe subcortical structures (hippocampus and amygdala). Unlike most DBS electrodes, which have stimulation contacts only at the tip, SEEG electrodes feature contacts along their entire length, enabling stimulation of multiple locations along their length. With an average of 8-10 electrodes, circa 70-100 contacts may be implanted per patient [10]. This configuration allows for recording and test stimulation at intended targets and incidental sites along the electrode trajectory.

While patients typically undergo telemetry evaluation normally lasting one or two weeks, the window for conducting recording and stimulation studies is often highly constrained by clinical and logistical limitations. Accordingly, exploring all contact combinations is impractical.

A viable solution involves identifying and prioritising the most informative combinations and testing as many as feasible within the practical constraints of each case. This approach requires careful optimisation to maximise the clinical and scientific insights gained from the limited testing opportunities.

Here we propose an initial approach to identifying such combinations of patients, contacts and tasks. Our approach leveraged the methodology well-established in the DBS field in the recent years and implemented in the Lead-DBS toolbox (https://www.lead-dbs.org/, (Neudorfer et al., 2023)). The main challenge we address is how to apply the methods developed for actual cases with known stimulation parameters and quantified clinical improvement to the scenario where only the implantation plan and the cortical regions involved in the clinical symptoms, or the task of interest are known. We also assume that stimulation effects on circuits implicated in psychiatric pathology can be observed in individuals without clinically significant psychiatric symptoms. This assumption is supported by recent advances in computational psychiatry [12] and the neurophysiology of psychiatric disorders [13–15]. Our analysis focuses on stimulation effects; however, circuit mapping could also leverage recordings of task-related activity or circuit-specific physiological biomarkers [16].

## Methods

Our analysis proceeded in two stages. At the first stage, we used data from previously published studies of DBS for OCD where the details of stimulation parameters and clinical improvement were available [2,17] to validate a predictive model not using these details. This involved two changes with respect to the previously published analyses. First, we replaced the biophysically informed Volume of Tissue Activated (VTA, [11]) with a generic spherical VTA that does not require stimulation parameters. Second, we replaced the similarity with empirical signature of improvement maps (R-maps, [18]) by similarity with meta-analytic activation map for OCD-related keywords obtained from the Neurosynth database (Figure 1a, https://neurosynth.org/, (Yarkoni et al., 2011)). We show that with these two modifications, we can still predict the clinical improvement well and obtain the optimal parameters for the predictive linear model.

**Figure 1:**
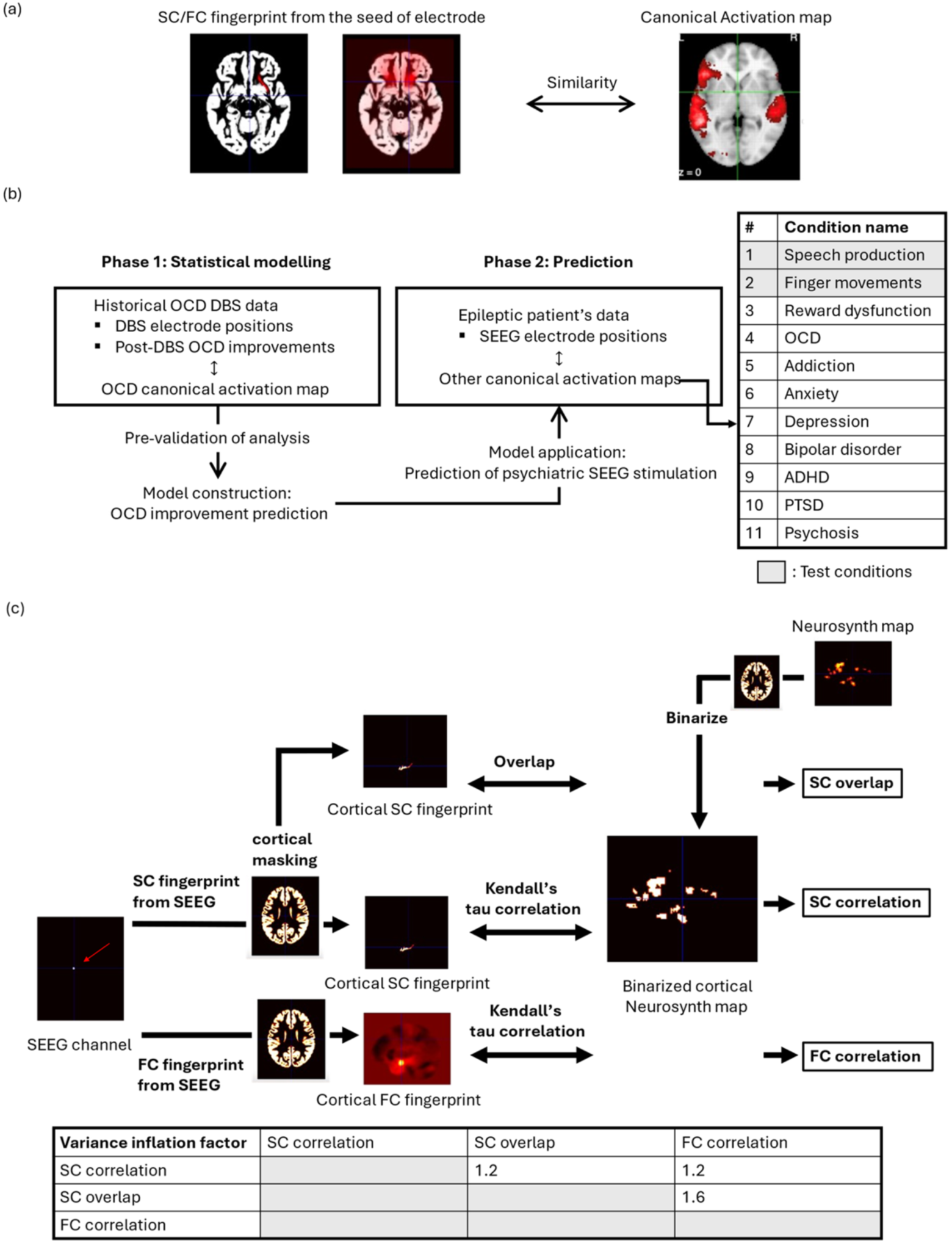
Overview of the analysis method. (a) Schematic representation of the proposed approach. Similarity between Neurosynth maps of psychiatric symptoms and connectivity fingerprints from SEEG contact VTAs is quantified and used to predict SEEG stimulation effects. (b) Two-stage study design: first, a statistical model is constructed using OCD DBS data; second, the model is applied to predict SEEG stimulation effects. The table lists the conditions analysed, with reference conditions (marked in grey) and psychiatric conditions of interest. (c) Similarity analysis pipeline for SC correlation, SC overlap, and FC correlation (top). SC and FC fingerprints, along with Neurosynth maps, are masked with a cortical template. SC overlap is calculated as the dot product between a Neurosynth map and a smoothed cortical SC fingerprint, normalised by the Neurosynth map area. SC correlation is determined using Kendall’s tau correlation between a Neurosynth map and a cortical SC fingerprint. FC correlation is computed similarly but with FC fingerprints. The variance inflation factor (bottom) demonstrates low collinearity among the three measures, indicating that they capture distinct connectivity properties.

In the second stage, we applied our predictive model to a large set of implantation plans from SEEG patients previously treated at the National Hospital for Neurology and Neurosurgery. The objective was to evaluate the number of electrode sites in these patients that could potentially serve as effective DBS targets for various psychiatric symptoms and related cognitive tasks. This analysis identified the symptoms and disorders most amenable to SEEG stimulation studies and estimated the patient numbers required for such investigations.

We first explain our model construction approach. We then describe the SEEG cohort analysis, and method of sample size estimation for SEEG predictive map construction based on the model. The complete workflow is summarized in Figure 1b.

## Model construction on DBS data

### Clinical data

The OCD dataset was acquired from four international DBS centres and used in previous publications [2,17]. DBS targets included bilateral Subthalamic Nucleus (STN), Nucleus Accumbens (NAcc) and Anterior Limb of Internal Capsule (ALIC). Locations of active DBS contacts in Montreal Neurological Institute (MNI) template space and the degree of improvement post-DBS measured with Yale-Brown Obsessive Compulsive Scale (Y-BOCS, [20]) for 80 patients were used for our model construction. The raw Y-BOCS scores were converted to relative improvement ([post DBS – pre DBS]/pre DBS) in line with the previous studies [2,17]. All patients gave written informed consent, and study protocols received ethical clearance from each local Ethics Committee.

### Canonical activation map preprocessing

Uniformity maps related to common psychiatric conditions (Table 1, Figure 1a) were acquired from Neurosynth (https://neurosynth.org/, [19]) and used as canonical activation maps in this study. All the maps were generated on 30/03/2024.

**Table 1.** Reference (grey) and psychiatric conditions with the corresponding keywords used to search in the Neurosynth site.

|  | Conditions | Keywords |
| --- | --- | --- |
| 1 | Speech production | 'speech production' |
| 2 | Finger movements | 'finger movements' |
| 3 | Reward circuit dysfunction | 'reward' |
| 4 | OCD | 'obsessive', 'compulsive', 'ocd' |
| 5 | Addiction | 'addiction' |
| 6 | Anxiety | 'anxiety' |
| 7 | Depression | 'depression', 'major depression' |
| 8 | Bipolar disorder | 'bipolar' |
| 9 | ADHD | 'adhd' |
| 10 | PTSD | 'posttraumatic', 'ptsd' |
| 11 | Psychosis | 'psychotic' |

“Finger movement” and “Speech production” (conditions 1 and 2) served as reference conditions expected to show strong effect predictions. This is because intracranial electrodes, placed to identify the epileptogenic zone, frequently traverse eloquent regions involved in language and sensorimotor functions. We also included ‘Reward circuit dysfunction’ as a construct hypothesised to contribute to most psychiatric disorders [21].

Binarized thresholded z-score maps from Neurosynth were warped from original MNI152linear template into MNI152NLin2009Asym template used in Lead-DBS toolbox using nearest-neighbour interpolation. When having several keywords related to the same psychiatric condition, as indicated in Table 1, we combined the corresponding binary maps by taking their union. The same procedure was applied to all conditions listed in Table 1.

### SC similarity analysis

A normative structural connectome, available within the Lead-DBS toolbox and derived from diffusion data of 32 subjects from the MGH-USC HCP dataset [22], was used to generate structural connectivity (SC) fingerprints.

These fingerprints were created at a 2mm x 2mm x 2mm resolution, matching the resolution of the Neurosynth maps. They were seeded from a 3.5mm sphere centred on the stimulating contact. This spherical volume of tissue activated (VTA) was used instead of a biophysical VTA, as the specific stimulation parameters required to generate a biophysical VTA in SEEG stimulation are yet to be determined. The 3.5mm spheres are similar in size to biophysical VTAs estimated for the OCD DBS cohort in the previous studies [2,17].

Two different analyses were used to assess similarity between the SC fingerprint and pre-processed Neurosynth maps: “SC correlation” and “SC overlap”. For SC correlation, non-cortical regions of the SC fingerprint and Neurosynth map were masked out. Since the pre-processed Neurosynth map was binary, Kendall’s tau correlation [23] was used to quantify similarity. For SC overlap, spatial intersection of voxels between the two maps (dot-product) was quantified and normalised by the number of suprathreshold voxels in the Neurosynth map.

### FC similarity analysis

FC similarity was calculated based on a normative functional connectome [24] derived using data from 1000 healthy subjects in the Brain Genomic Superstruct Project [25,26]. Unlike SC, FC fingerprints contain both positive and negative values. Positive FC reflects regions whose signals fluctuate in the same direction as the seed region, while negative FC reflects regions with opposing signal patterns. This makes direct comparison with non-negative Neurosynth maps less straightforward. Nevertheless, when the positive and negative components of FC were examined separately, both showed positive associations with clinical improvement. Although seemingly contradictory, this arises from our use of Kendall’s tau correlation, which ranks values based on their order without considering their sign or magnitude. As a result, separating FC into positive and negative subsets yields highly similar rank structures and correlations with outcome, introducing redundancy. These findings suggest that the strength of connectivity—regardless of its direction—is more relevant to clinical response.

One might consider using the absolute value of FC to capture this effect. However, because Kendall’s tau relies only on the rank order of values, taking absolute values removes information that is already discarded by the rank transformation, thus disrupting the relationship entirely. For consistency and simplicity, we therefore applied the same correlation procedure used for SC, but with the FC fingerprint. The resulting measure is referred to as the ‘FC correlation’.

### Predictive model

For multivariate linear regression model based on the SC correlation, SC overlap, and FC correlation, it is essential to assess the collinearity between these variables. The collinearity was analysed using the variance inflation factor, which indicates higher collinearity with higher values (Figure 1c).

A multiple linear regression model was developed to predict improvements following DBS treatment. To make the model coefficients comparable, SC overlap and FC correlation were normalised by multiplication with the ratio of the mean SC correlation to the mean values prior to fitting the model. The predictive power of the approach was verified using leave-one-out cross-validation for OCD DBS data. Finally, the model from full set of OCD DBS data was constructed as follows (Figure 2):

**Figure 2:**
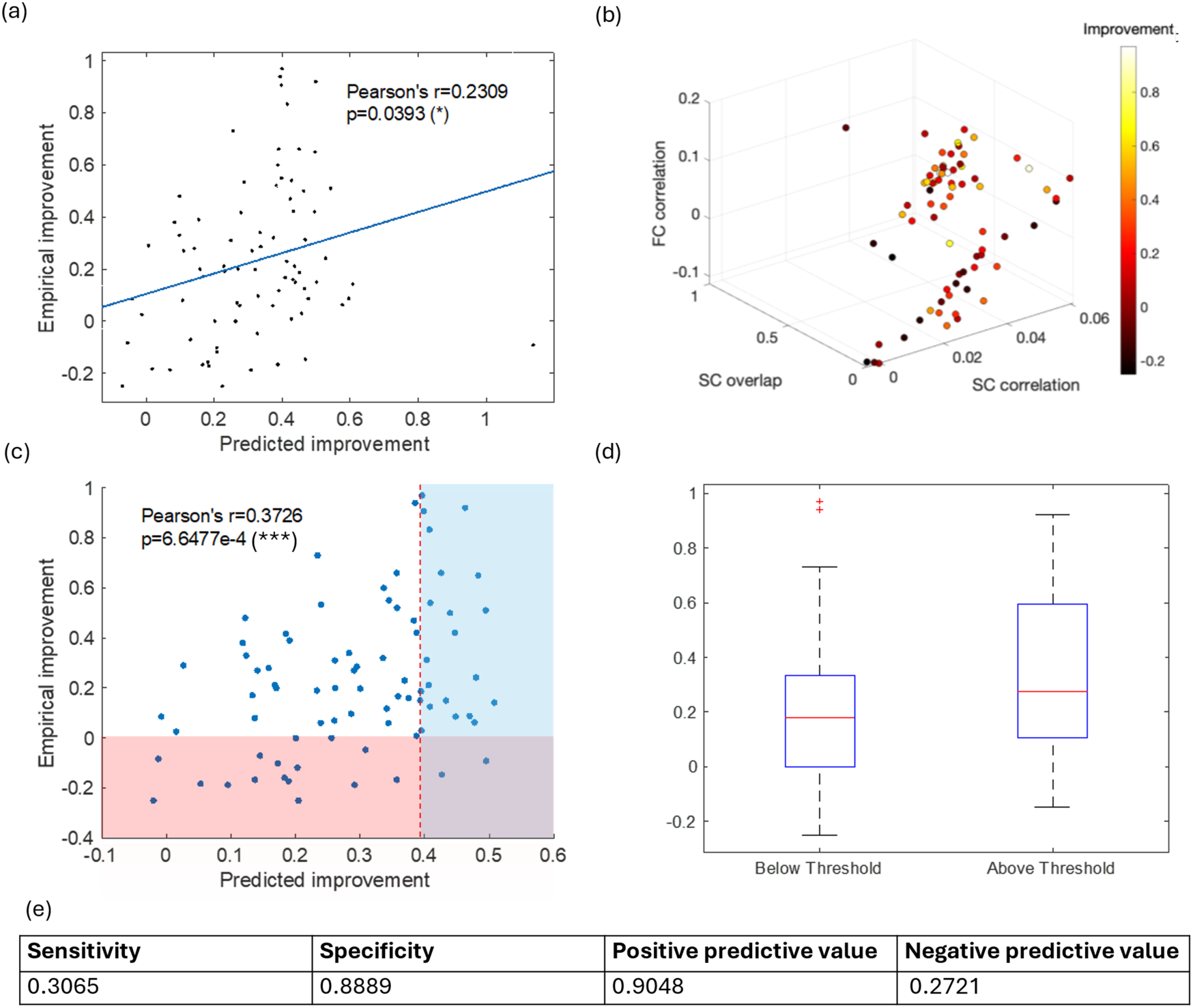
Predictive power of the linear model for OCD DBS data. (a) Leave-one-out cross-validation demonstrating significant predictive power of the model in OCD DBS data. The light blue shading represents the 95% confidence interval for the regression of predicted versus empirical improvement. (b) 3D plot illustrating the relationship between SC correlation, SC overlap, FC correlation, and clinical improvement, with improvement represented by colour. (c) Thresholding the predictive score. The red dashed line marks the lower threshold for predicted improvement, set at the 90th percentile of cases with no or negative empirical improvement, ensuring that predicted improvement corresponds to positive empirical improvement. (d) Box plot comparing groups with predicted improvement below and above the threshold (top), alongside the model’s performance in OCD DBS data (bottom). The interquartile range of the “below threshold” group includes zero and negative empirical improvement, whereas the “above threshold” group excludes them, supporting the validity of the thresholding approach.

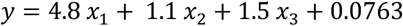

where *x*_1_ is SC correlation, *x*_2_ is SC overlap, *x*_3_ is FC correlation, and *y* is post-DBS improvement.

To identify contacts where stimulation could potentially influence symptoms, the model’s predictive scores were thresholded. The threshold was defined as the point at which the model achieved a positive predictive value greater than 0.9 and specificity greater than 0.85 (Figure 2c). Scores exceeding this threshold were considered predictive of empirical effects.

## Model application to SEEG data

### Clinical data

For this analysis, we used locations of SEEG electrode contacts of 86 patients with drug-resistant, focal epilepsy that underwent invasive recordings at the National Hospital for Neurology and Neurosurgery, Queen Square, UK. The total number of electrode contacts in all patients was 6372 with 6-10 contacts in an electrode and 16-111 contacts in a patient.

We automatically identified the location of SEEG contacts in native space following [27], a process that was manually verified. For each SEEG electrode, we extracted contact locations and transformed their coordinates into MNI space. The transformation consisted in applying a rigid registration first, followed by an affine registration, and then a nonlinear (F3D) registration, whereby the skull-stripped T1 image in native space was used as the starting floating image and the skull-stripped MNI image (0.5 mm x 0.5 mm x 0.5 mm) was used as a reference. The distribution of the spherical VTAs corresponding to all the contacts is shown in Supplementary Figure 1.

### Similarity analysis and prediction

SC and FC connectivity fingerprints from all SEEG electrode contacts were generated as described above. The SC correlation, SC overlap, and FC correlation values between connectivity fingerprints from each electrode contact and Neurosynth maps for different test and psychiatric conditions (Table 1) were computed. The model constructed from OCD DBS data was applied to the three similarity measures to predict the magnitude of SEEG stimulation effects. Predictions were binarised into ‘effect’ or ‘no effect’ using a threshold derived from the OCD DBS dataset.

### Sample size estimation

We next aimed to estimate the number of SEEG patients that would have to be tested in order to derive a symptom-specific map for predicting stimulation effects using the methodology previously established for DBS. To identify brain networks correlated with clinical improvement induced by DBS, the correlation coefficients between network maps and the post-DBS improvement can be presented as an R-map [18]. The required sample size for building such a map *N*can be estimated as

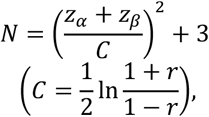

where *r* is the expected correlation coefficient, *z_α_* is Type 1 error, and *z_β_* is Type II error [28]. We took 0.5 as the expected correlation coefficient based on the peak values of previously published R-maps [17,18]. Assuming Type I error of 0.05 and a Type II error of 0.20, it gives 29 measurements necessary. To calculate the sample size in terms of patient number, we divided this number by the mean number of effective contacts per patient derived from our previous analysis. The upper range was calculated with 95% confidence interval of the number of effective electrodes. The lower range was division of 29 by the probability to have at least one effective contacts for the psychiatric condition.

## Results

### Model validation on DBS data

SC correlation, SC overlap, and FC correlation were all significantly correlated with OCD improvement after DBS (Spearman’s r=0.27, p=0.017; r=0.36, p=0.0009; r=0.25, p=0.024). Low variance inflation factors of the three pairs of two measures (Figure 1c) suggested that the three measures were linearly independent of each other, so they could be combined in a multiple regression model.

We, therefore, combined the three measures in a linear model to provide a single predictive score (see Methods). Scores exceeding a threshold defined as the point at which the model achieved a positive predictive value greater than 0.9 and specificity greater than 0.85 were considered predictive of empirical effects. Indeed, patients in the above-threshold group showed significantly greater improvement than those in the below-threshold group (one-tailed Mann–Whitney test, p = 0.0256). The performance of this method on OCD DBS data is shown in Figure 2e. Note that in the case of OCD, the observed effects were symptom improvements due to the way targets were selected. However, for other target–symptom combinations, stimulation could also result in symptom worsening.

The predictive power of the model based on the three measures was validated with leave-one-out cross-validation in Figure 2a (Pearson’s r=0.23, p=0.039). For the group with predicted improvement above the threshold of 0.3325 (red dashed line in Figure 2c), the interquartile range of empirical improvement did not include zero (Figure 2d), confirming that thresholding of predicted improvement distinguished the group with positive empirical improvement from the entire cohort of OCD DBS data in Figure 2b. The sensitivity, specificity, positive predictive value, and negative predictive value of the model are provided in Figure 2e. As the threshold is set for high positive predictive value, while it gives low performance for other tests, positive predictive value and specificity are close to 90%.

A potential concern is whether the similarity scores and predictive model are condition-specific. To address this, we tested whether similarity scores between SC fingerprints and non-OCD Neurosynth maps correlate with OCD symptom improvement (Supplementary Figure 2a). The results show that SC correlations exhibit condition specificity: only OCD-related SC correlations significantly and positively correlate with OCD improvements, whereas SC correlations for other conditions show either non-significant or negative correlations. In contrast, FC correlations and SC overlaps do not demonstrate condition specificity, as FC correlations and SC overlaps derived from other Neurosynth maps also significantly correlate with OCD improvements.

We then tested the predicted effects of DBS in OCD targets on other conditions using Neurosynth maps and our model. This analysis showed that, after OCD, the highest predicted effects were observed in Reward Dysfunction, Addiction, and ADHD (Supplementary Figure 2c). The variation in predictions across conditions could be due to their relatedness to OCD or differences in the number of suprathreshold voxels in their respective Neurosynth maps. Further analysis shown in Supplementary Figure 2d demonstrated that the differences in predictions across conditions were strongly driven by the cross-correlation of each condition’s Neurosynth map with the OCD Neurosynth map (Spearman’s r = 0.8737, p < 0.001). However, there was no significant correlation with the total number of active voxels in the Neurosynth maps (Spearman’s r = 0.2067, p = 0.5420).

### Predicting SEEG stimulation effects

Figure 3a and 3b show the similarity measures and predictions of stimulation effects for each SEEG contact and all the conditions in a single representative patient. The three similarity matrices show different patterns depending on the analysis method. The network expected to be most affected by stimulation in this patient is ‘Reward dysfunction,’ and the contact most expected to affect the task performance related to reward is contact 33 in right inferior frontal gyrus based on Automated Anatomical Labeling Atlas 3 [29]. Finger movement, speech production, reward dysfunction, and bipolar disorder could be affected with stimulation of at least one SEEG contact in this patient.

Model predictions across the whole SEEG cohort are summarised in Figure 3c. This matrix shows the patients for which there was at least one suprathreshold contact for the corresponding condition. The reference conditions, finger movement and speech production, are predicted to be commonly affected with SEEG stimulation for over 80% of patients. Among the psychiatric conditions, reward dysfunction could be affected in over 80% of patients, followed by depression and bipolar disorder with over 50% of patients predicted to be affected.

**Figure 3:**
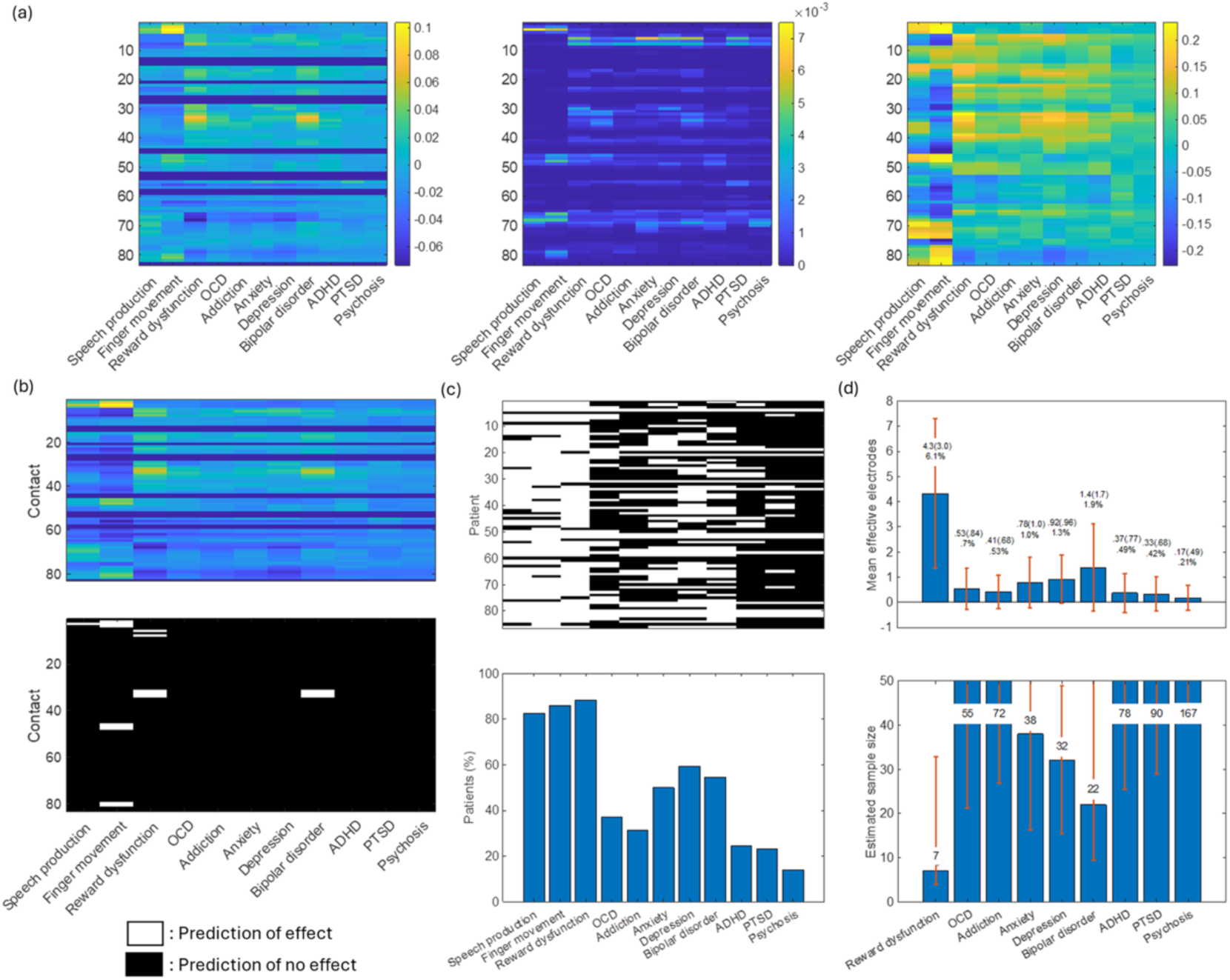
Summary of stimulation effects prediction for a representative patient and all patients. (a) Matrix displaying similarity scores for each feature: SC correlation (left), SC overlap (middle), and FC correlation (right). (b) Predicted stimulation effects for a single patient. The top matrix shows the predicted effect magnitude for all conditions and electrode contacts. The bottom matrix presents thresholded predictions, where white indicates effects above the threshold and black indicates effects below. Predictions suggest stimulation may influence speech production, finger movement, reward dysfunction, and bipolar disorder in this patient. (c) Group-level predictions across all patients. The top matrix displays maximal thresholded effect predictions per condition, with the x-axis representing conditions and the y-axis representing patients. The bottom bar plot indicates the percentage of patients predicted to have at least one electrode contact with an effect above the threshold. Tasks related to finger movement, speech production, and reward dysfunction are predicted to be affected in most patients. (d) The top bar plot shows the number of effective electrode contacts for psychiatric conditions per patient. The upper row of numbers above each bar represents the mean number of effective contacts and its standard deviation (in parentheses), while the lower row provides the mean percentage. Error bars indicate standard deviations. The bottom bar plot presents the estimated required sample size for studying psychiatric networks using SEEG stimulation, calculated by dividing the target of 29 effective electrode contacts by the mean number of effective contacts per patient. Conditions other than reward dysfunction use an alternative method for the upper error bar, based on the percentage of patients predicted to have at least one effective contact. Tasks related to reward dysfunction are expected to require the smallest sample size.

The top bar plot of Figure 3d shows the mean number and percentage of electrode contacts predicted to affect each symptom. The reward dysfunction has the highest mean number of effective contacts per patient.

The mean number of effective electrode contacts per patient can also be used to estimate the number of participants required to construct a predictive map of SEEG stimulation effects (R-map). Assuming a maximum Pearson’s correlation coefficient (r) of 0.5, derived from the highest R-value in causal R-maps, for the potential maximal R-value of SEEG R-map, the required minimum number of SEEG stimulation data points is 29. This sample size refers to the number of effective electrode contacts, not the number of patients, since a single epilepsy patient could have multiple effective SEEG contacts. The corresponding numbers of patients are shown at the bottom of Figure 3d. The condition requiring the least number of patients is ‘reward dysfunction’ with sample size of 7. Even in the worst case assuming effective contact count of one standard deviation below the mean, a study of dysfunctional reward processing is expected to be accommodated with 33 patients. It is followed by ‘bipolar disorder’, ‘depression’, and ‘anxiety’ with 22, 32, and 38 patients respectively. For other psychiatric conditions, sample size is estimated to be over 40, making such studies impractical at least in a single centre. Stimulation studies could be made more efficient by using normative maps like Neurosynth as a starting point and iteratively updating them with each measurement. This adaptive approach, which is yet to be developed, could refine the map toward the desired R-map, following the principles of active sensing [30]. To obtain an upper bound on the difference such an approach could make, we repeated the calculation for OCD using the FC correlation with OCD DBS FC R-map instead of an OCD Neurosynth map, keeping other similarity features in the model the same (Supplementary Figure 3). Using this map would reduce the number of SEEG patients required to re-estimate it by 78% (from 55 to 12).

## Discussion

We evaluated the potential of intracranial stimulation of SEEG patients as a platform for testing novel DBS targets. Since SEEG electrodes in patients with epilepsy are used to map the epileptogenic zone, it remains uncertain whether they can effectively target psychiatric networks. Our results show that for disorders of reward (such as anhedonia) and for bipolar disorder this could be a promising approach. We must reiterate, however, that we only provide a feasibility study and our conclusions are based on the implantation plans from one clinical site and could well be different for other sites. Furthermore, although the ranking of circuits by the estimated number of patients required for a study was consistent across multiple analytical variants, the absolute patient numbers were sensitive to specific choices in the processing pipeline. We, therefore, anticipate that future studies could revise these estimates, potentially in either a more optimistic or more conservative direction.

### Future work

Future work can potentially refine our methods in several different ways. We used the classical version of Neurosynth and a simple keyword-based meta-analysis. The more recent version ‘Neurosynth Compose’ (https://compose.neurosynth.org/, [31]) makes it possible to make a fine-grained selection of relevant studies and generate more specific meta-analytic maps. Moreover, for studies of stimulation effects on specific tasks, maps from fMRI studies of these tasks can be used instead.

Task-based functions (i.e. motor control, language) may represent an ideal platform to refine and validate the proposed method [32,33]. For the tasks such as speech production and finger movement, the estimated sample sizes required to achieve a significant empirical effect in a t-test comparing groups with predicted effect versus no effect would be approximately five and four patients, respectively. We predict this should be easily achieved considered the contact distribution in our cohort. The calculation assumes mean rates of effective contacts per patient of 6.1% and 9.6%, a Type I error rate of 0.05, and a Type II error rate of 0.20 [28,34]. The standard deviation (0.3) and mean difference in empirical improvement (0.19) between subthreshold and suprathreshold groups in the OCD DBS dataset were used for these estimates.

Anhedonia affects up to 50% of people with epilepsy (Roberts-West et al., 2023), and therefore clarifying its underlying network is particularly relevant for this group. Our results suggest that anhedonia circuits may often be transversed by SEEG contacts, making the delineation of this circuit via stimulation a priority. However, as mood-related effects may not manifest immediately or may be challenging to classify and quantify in real time, task-based assessments susceptible to disruption by stimulation could provide alternative indicators of therapeutic potential. Developing such tasks is a key focus in computational psychiatry, a field recently proposed to benefit from joining forces with DBS and intracranial monitoring [12]. The availability of validated tasks could also remove the constraint of studying only patients with comorbidities. Task development and validation should therefore progress alongside further refinement of stimulation techniques and network characterisation.

Another important research direction is the identification of neurophysiological biomarkers for psychiatric symptoms. Recent breakthroughs in this field [13–15,36] suggest that testing the effects of stimulation on such biomarkers could provide an alternative to behavioural assessments. These biomarkers could be derived from both SEEG recordings and non-invasive concurrent high-resoluton scalp EEG, offering additional methods to evaluate stimulation effects.

Both computational psychiatry and biomarker research have strong links to animal studies. Mapping homologous circuits between humans and animals, and establishing forward and reverse translation in stimulation research, could further facilitate the identification of effective DBS targets using SEEG.

In our approach, the predictive score for each contact is fixed throughout the study. This could be improved by using the initial, map-based prediction only as a prior, updating the scores with each new result and incorporating the informativeness of testing each contact, rather than its potential effectiveness. For instance, if a particular site has already been tested several times, other sites should be prioritised in the future. Such a framework would be akin to ‘active sensing’ proposed for other applications [30], and we showed that, in the best-case scenario, it could reduce the required patient number by 78% (Supplementary Figure 3), with a similar approach applied to SC potentially affording further improvement.

### Limitations

As a retrospective study, our predictive analysis has several limitations. Strictly speaking, the model constructed using OCD DBS electrodes cannot be directly generalised to SEEG electrodes. While OCD DBS contacts are confined to a narrow region around the subthalamic nucleus and the anterior limb of the internal capsule, SEEG contacts are more widely distributed across the hemisphere and specific target cortical regions. However, our key assumption was that the proportional relationship between connectivity scores and stimulation effects would be preserved.

Another potential issue is that SEEG leads are considerably thinner than DBS leads, resulting in a smaller electrode surface area. This increases the local charge density adjacent to the electrode while producing a smaller VTA. Therefore, SEEG stimulation parameters required to achieve effects comparable to those of DBS may need to be systematically determined. Recent advances in VTA computational modelling [37] now allow much of this work to be conducted *in silico* prior to patient testing.

Of the three similarity scores used, only SC correlation exhibited high condition specificity. However, leave-one-out cross-validation of a linear model based solely on this measure showed no significant predictive power. In contrast, a model incorporating all three similarity scores—SC correlation, SC overlap, and FC correlation— demonstrated significant predictive power (Figure 2a). These measures capture distinct aspects of connectivity with low collinearity (Figure 1c).

SC correlations reflect the spatial similarity between regions connected by SC fingerprints and activated regions, independent of magnitude, and are not influenced by the VTA’s relative proximity to white matter. SC overlap quantifies proximity to effective fibre tracts. FC correlations capture the spatial similarity between FC fingerprints and activated regions, differing from SC correlations due to their inclusion of indirect connectivity beyond white matter tracts, as indicated by their low collinearity (Figure 1c).

While individual similarity scores lack complete condition specificity, their combined use enhances predictive accuracy, resulting in a more robust model. Absolute specificity may not be expected, as Neurosynth patterns and the corresponding networks involved in psychiatric symptoms often overlap. Furthermore, the finite number of distinct FC networks in the brain [38] inherently limits the potential specificity of FC correlation.

The choice between uniformity and association Neurosynth maps also requires consideration. The uniformity test assumes activation is evenly distributed across the brain, while the association test compares activation in condition-related and unrelated regions. Previous studies have used association maps to assess overlaps with FC fingerprints in DBS research [39], while others have used uniformity maps to compare OCD DBS R-maps with Neurosynth maps for non-invasive stimulation applications [17]

In our OCD dataset, the association map covered too small an area for statistical analysis, with only nine of eighty patients showing a non-zero VTA overlap. While SC overlap with the association map correlated with improvement, FC correlation did not. Uniformity maps, covering broader activation regions, better represent networks rather than localised associations. Since DBS primarily targets networks, uniformity maps are more suitable for our analysis. Their larger activation areas also make them preferable for quantitative comparisons of FC fingerprints and Neurosynth maps.

A potential issue with research in people with epilepsy is that severe epilepsy can induce functional reorganisation of the brain [40,41]. As our analysis focuses on psychiatric networks in the general population, and assuming there are no distinct structural connectivity changes in patients with psychiatric illness, we used a normative connectome and Neurosynth maps without taking into account epilepsy-related alterations. This is unavoidable when joining different patients in a normative space. We believe that the issue of atypical anatomy can be addressed by excluding patients with known developmental abnormalities and avoiding stimulation sites near epileptogenic tissue. Additionally, for both clinical and ethical reasons, stimulation near suspected epileptogenic zones would not be conducted for general research purposes. Exclusion of some fraction of potentially suitable contacts would increase the number of patients required to complete a study.

Our findings suggest that the predictive model developed for OCD DBS may have broader applicability beyond OCD, as conditions with higher predicted effects were those most closely related to OCD rather than those with larger activation volumes in Neurosynth maps. The strong correlation between prediction differences and cross-correlations of Neurosynth maps (Spearman’s r = 0.87, p < 0.001) supports the notion that the model captures meaningful connectivity patterns associated with psychiatric symptoms. However, the greater variance observed in SEEG predictions across conditions (Figure 2c) compared to OCD DBS predictions (Supplementary Figure 2c) suggests key differences in electrode placement and the underlying connections. While connections passing via the ALIC are often crossed by electrodes in our SEEG sample, SEEG electrodes also frequently target more superficial white matter tracts with functionally distinct roles, potentially reducing the generalisability of DBS-based predictions to SEEG stimulation. Despite these differences, the model’s ability to predict stimulation effects for related disorders highlights its potential utility in guiding target selection for future research. Direct validation of the model’s applicability to SEEG stimulation remains necessary to confirm its predictive value in this distinct neuromodulatory context.

## Conclusion

This study explores the feasibility of using intracranial monitoring in epilepsy as a platform for developing novel DBS targets and mapping circuits underlying psychiatric symptoms. Our results indicate that impairments related to reward processing, such as anhedonia, and mood disorders are the most promising syndromes for investigation. The methods presented here can be readily applied to different implantation plans and alternative approaches to circuit characterisation, enabling more refined and personalised stimulation protocols.

## Acknowledgements

This research did not receive any specific grant from funding agencies in the public, commercial, or not-for-profit sectors. HA is supported by the National Institute for Health and Care Research University College London Hospitals Biomedical Research Centre. DG is supported by the Epilepsy Research Institute UK with an Emerging Leaders fellowship (F2403).

## Declaration of generative AI and AI-assisted technologies in the writing process

During the preparation of this work the authors used ChatGPT for language editing of human-written text. After using this tool, the authors reviewed and edited the content as needed and take full responsibility for the content of the published article.

## Code and data availability

The code used in this study is available at: https://github.com/vlitvak/SEEG_DBS. Data can be shared upon reasonable request. For access to the OCD dataset, contact NL; for the SEEG dataset, contact DG.

## Author Contributions

Haun Sun: Formal analysis, Visualization, Writing – original draft; Ningfei Li: Conceptualization, Software, Writing - Review & Editing; Alejandro Granados Martinez: Formal analysis; Fahmida A. Chowdhury: Investigation; Beate Diehl: Investigation, Writing - Review & Editing; Harith Akram: Investigation, Writing - Review & Editing; Veerle Visser-Vandewalle: Investigation, Writing - Review & Editing; Bryan Strange: Investigation, Writing - Review & Editing; Juan A. Barcia: Investigation; Mircea Polosan: Investigation, Writing - Review & Editing; Stéphan Chabardes: Investigation, Writing - Review & Editing; Davide Giampiccolo: Conceptualization, Investigation, Data curation, Supervision, Visualization, Writing – review and editing; Vladimir Litvak: Conceptualization, Supervision, Writing – original draft

---

**Supplementary Figure 1:**
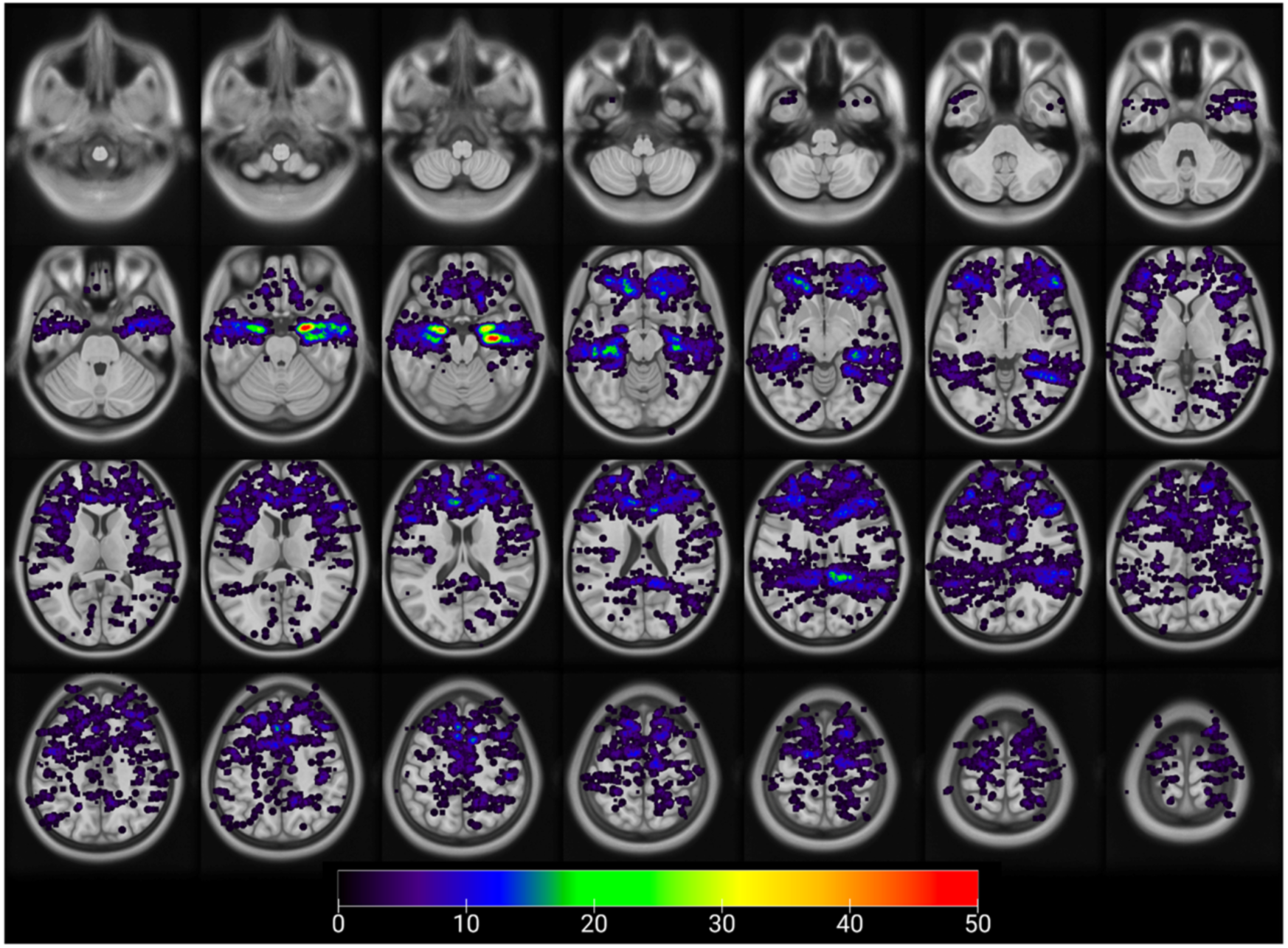
Distribution of spherical Volumes of Tissue Activated (sVTA) in the brain in our epilepsy cohort. The total number of electrode contacts in all patients was 6372 with 6-10 contacts in an electrode and 16-111 contacts in a patient. An in-house Python script was used to create on overlap map of all the 3.5 mm sVTAs in the MNI space. This was visualised in MRcroGL (https://www.nitrc.org/projects/mricrogl). The colour encodes the number of overlapping sVTAs in one location.

**Supplementary Figure 2:**
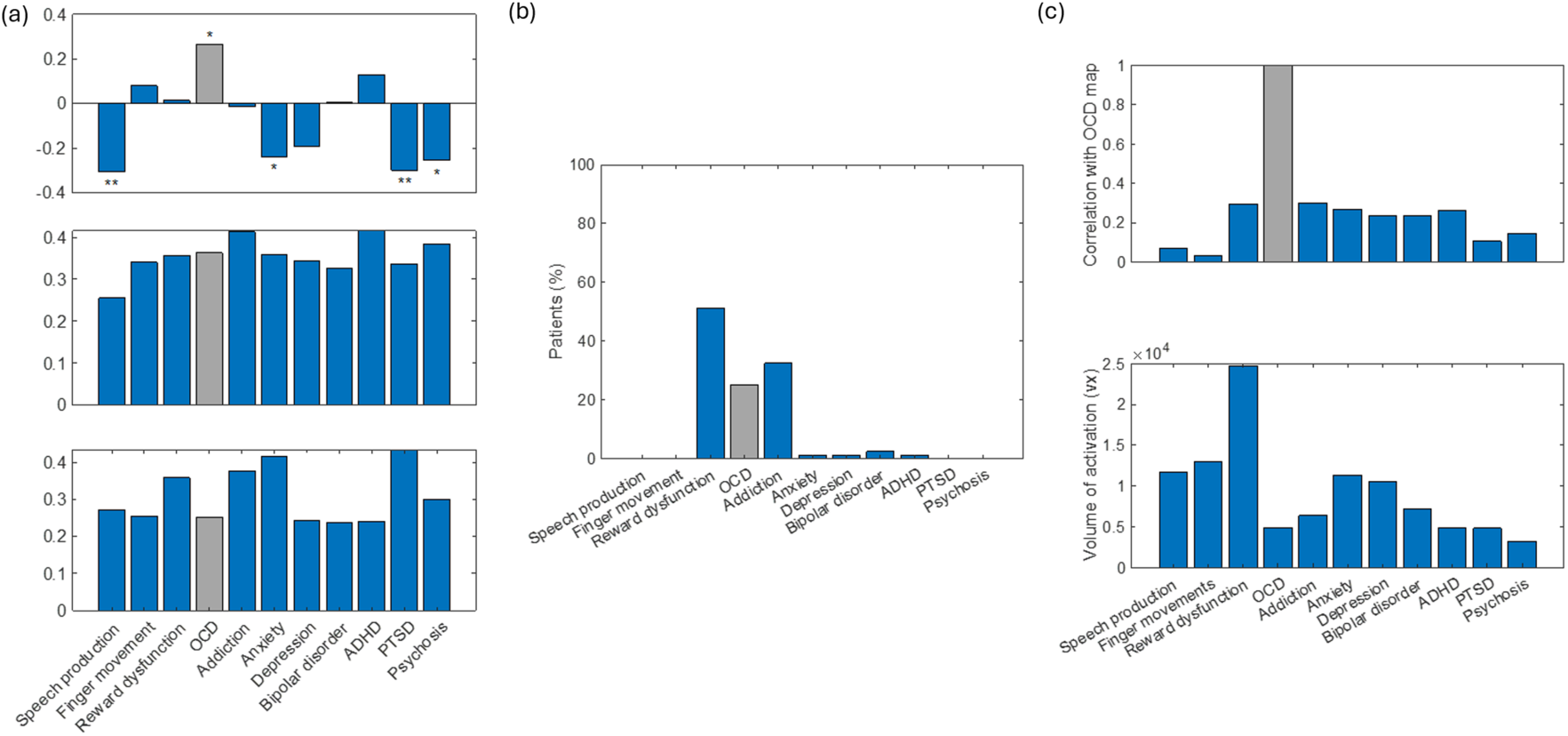
Condition-specificity of similarity scores and generalisability of the model beyond OCD. (a) Condition-specificity of SC correlation (top), FC correlation (middle), and SC overlap (bottom). The grey bar represents the correlation between OCD similarity scores and OCD improvement, while all other bars represent control data, showing correlations between similarity scores of other conditions and OCD improvement. Only SC correlations demonstrate a condition-specific, significant positive correlation with OCD improvement, whereas control data for FC correlations and SC overlaps also show significant correlations. (b) Prediction of the proportion of OCD patients affected by OCD DBS in tasks related to other conditions. Compared to SEEG stimulation predictions (Figure 3), the variance is generally lower. Following OCD, the highest predicted proportions of afected patients are seen in Reward Dysfunction, Addiction, and ADHD. (c) Relationship between OCD Neurosynth maps and other conditions. The top panel shows cross-correlation between the OCD Neurosynth map and maps of other conditions, while the bottom panel displays the volume of activation in Neurosynth maps. Cross-correlation, which reflects relatedness to OCD, is strongly correlated with prediction on control data (panel b), whereas activation volume is not. This suggests that the OCD model is generalisable to other conditions, with the predicted improvement following OCD DBS being linked to the condition’s similarity to OCD.

**Supplementary Figure 3:**
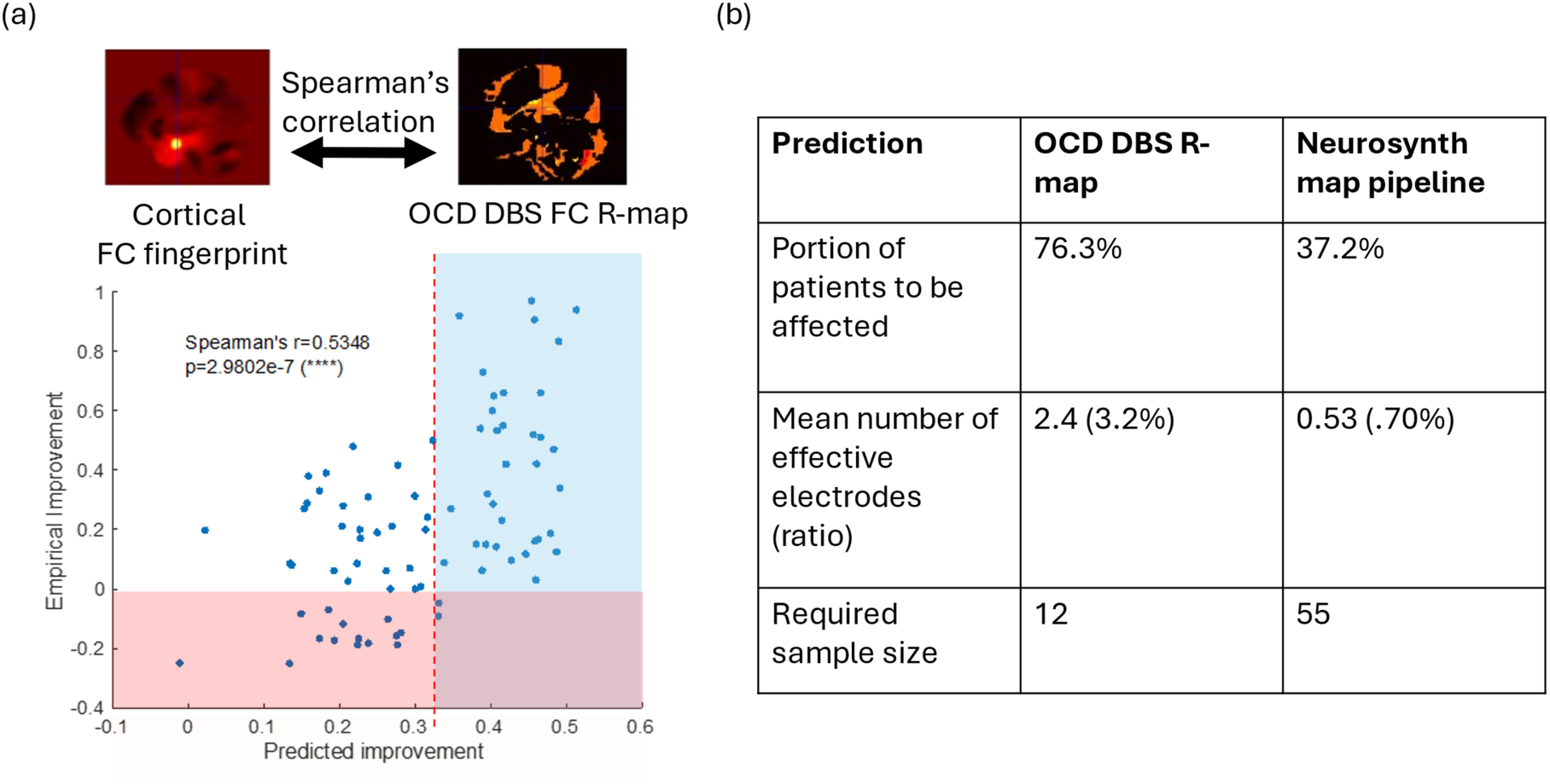
Prediction of SEEG stimulation effect on the OCD network using the OCD DBS FC R-Map for validation. (a) Methodology for calculating FC correlation with the OCD DBS FC R-map (top) and scatter plot of predicted improvement using the R-map pipeline, replacing FC correlations with correlations between FC fingerprints and the OCD DBS R-map (bottom, cf. Fig, 2C). To ensure a reliable empirical effect, the threshold was set at the 90th percentile of predicted values within the group showing no or negative improvement (0.30). (b) Predictions of SEEG stimulation effects on the OCD network in epilepsy patients, based on this threshold and similarity features. These predictions identify a greater number of relevant patients and electrodes than those obtained using the Neurosynth map, suggesting a high false-negative rate in the Neurosynth-based pipeline. This highlights the potential for an iterative approach, integrating the construction of an SEEG R-map following the initial Neurosynth-based prediction.

